# Effects of an Innovative Training Program for New Graduate Registered Nurses: a Comparison Study

**DOI:** 10.1101/2020.09.11.20192468

**Authors:** Fengqin Xu, Yinhe Wang, Liang Ma, Jiang Yu, Dandan Li, Guohui Zhou, Yuzi Xu, Hailin Zhang, Yang Cao

## Abstract

New graduate registered nurses (NGRNs) face great challenge during the transition from school to clinical practice. The aim of the study is to evaluate the effects of a new training mode for newly recruited NGRNs in a Chinese teaching hospital. This is a quasi-randomized controlled study. One hundred and fifty NGRNs were randomly selected from a teaching hospital and assigned into two groups. The conventional training and a new training program were taken for the control group and the research group respectively. At the end of training, the two groups were evaluated and compared for theoretical knowledge and operation skills using a self-assessment questionnaire and the Chinese Registered Nurse Core Competency Scale. The theoretical knowledge (88.4 vs. 81.7, p< 0.001), operation skills (94.8 vs. 90.3, p< 0.001), and total core competencies scores (156.2 vs. 148.8, p< 0.05) in the research group were significantly higher than those in the control group. Compared to the control group, the research group had statistically significantly higher scores in education and consultation (2.47 vs. 2.40), clinical nursing (2.87 vs. 2.62), interpersonal relationship (2.56 vs. 2.43), and critical thinking and scientific research (2.78 vs. 2.61). The innovative pre-job training program for NGRNs conducted in Chinese clinical nursing skill training bases could significantly improve the training effect and is worthy of broader implementation.

## Introduction

New graduate registered nurses (NGRNs) are the nurses who have work experience less than one year after registration. Researches indicated that NGRNs had lower capability to incorporate their knowledge, skills, and evidence-based practice into their work [1]. New nurses needed support and training to speed their transition from new graduates to qualified practitioners, and improve their retention [2]. Standardized training is important to NGRNs to complete this transition between school education and clinical nursing work.

Different countries and regions have different emphasis on training NGRNs. In the United States (U.S.), the NGRNs transition programs use the curriculum of the University Health System Consortium or the American Association of Colleges of Nursing Nurse Residency, which was developed by the Versant New Graduate Registered Nurses Residency [3]. The U.S. training mode is tailored according to the personal training needs of hospitals, NGRNs, and the reregistration of nurses. NGRNs can take part in academic conferences, seminars, lectures, and/or online courses, et al. They can also choose free continuing education programs that are approved by authoritative agencies or institutions and provided by hospitals for online courses training [4]. NGRNs learn how to develop nursing thinking through clinical practices of caring for a variety of patients, help and assistance from colleagues, and discussion with peers [5]. The training focuses on enabling NGRNs to learn relevant professional knowledge and expertise according to their individual expectations and to develop into a specific direction towards specialist nurses. In Canada, NGRN education mainly focuses on practical skill development [6]. In the United Kingdom (U.K.), the Flying Start of the National Health Service (NHS) is adopted as a national development program for all newly qualified nurses [7]. The main training contents of the NHS Flying Start are clinical ability, decision-making ability, teamwork, leadership and management ability, and career development planning, et al [8]. NGRNs can choose different training courses according to their own professional speciality and career development plan. They may adopt the model of combining training from both hospitals and universities.

In China, the conventional training for the graduate registered nurses pays more attention to the cultivation of theory and skills. The NGRNs face great challenge during the transition from the school to clinical practice. The transition from nursing student to practicing nurse can be very stressful for some new nurses. In Japan, the Transition Program for Newly Graduated Nurses is widely used [9]. The contents of this NGRN training contents include the clinical knowledge and skills required in a handbook of basic nursing procedures. Japan adopts collective education and on-the-spot education to train new nurses. The on-the-spot education is organized and implemented by various departments [10]. At the same time, NGRNs are encouraged to acquire confidence and strengthen professional identity through exchange experience [9, 11]. Although many healthcare organizations have tried to provide transition programs to support NGRNs through this vulnerable period, the optimum length and structure of the programs are still under exploration [12]. In general, the theoretical knowledge and clinical practice of the NGRNs is not closely connected, and there is still an unneglectable gap between clinical thinking and practical ability and the needs of daily clinical work.

To evaluate the effects of a new training mode for NGRNs, we conducted a quasi-randomized controlled study of NGRN training and compared two training modes in the newly recruited NGRNs in a tertiary and teaching hospital in Lianyungang City, Jiangsu Province, China. The main outcomes of interest were theoretical knowledge, operation skills, and core competencies of the NGRNs.

## Methods

### Participants and study setting

The study was approved by the Ethical Committee of the First People’s Hospital of Lianyungang (approval number: KY20171101001), and conducted between July 2018 to November 2018. Conducting of the study followed the Transparent Reporting of Evaluations with Nonrandomized Designs (TREND) statement [13].

To detect a statistically significant difference of 3 points between the mean scores of the two groups, with a type I error of 0.05, power of 0.9, and standard deviation of 5, 63 participants were needed for each group. Considering 10 drop off rate, one hundred and fifty NGRNs (7 males, 143 females) were randomly selected from the First People’s Hospital of Lianyungang. They were randomly assigned into the research and control groups using their registration numbers. Each group had 75 NGRNs. All the NGRNs participated n the pre-job training in July 2018 immediately after they were recruited by the hospital.

Trainings were implemented in the Clinical Nursing Skills Training Base (CNSTB) of the hospital. The required nursing knowledge and skills in the CNSTB were obtained and evaluated by the nursing department in hospital. The goals of CNSTB are: 1) to facilitate transition of new graduate nurses to professional registered nurses (RNs); 2) to prepare an entry level staff nurse with confidence who can provide patients with quality and satisfactory care; and 3) to increase the commitment and retention of new graduate nurses within the organization [14].

Immediately after the NGRNs’ were recruited, they participated in a 4-week pre-job training in the CNSTB organized by the Department of Human Resources (DHR) and the Department of Nursing (DN) of the hospital. The training in the first week was the new staff training organized by the DHR. The training from the second to the fourth week, including theoretical knowledge and operational skills, was organized by the DN and eight hours per day.

The training forms included in-classroom teaching, case analysis, group discussion, operation demonstration, and scenario simulation, et al. All of the contents were designed to improve the effects of new nurse training.: The enrollment, assignment, allocation, follow-up, and analysis of the participants are show in Figure 1.

**Figure 1.**
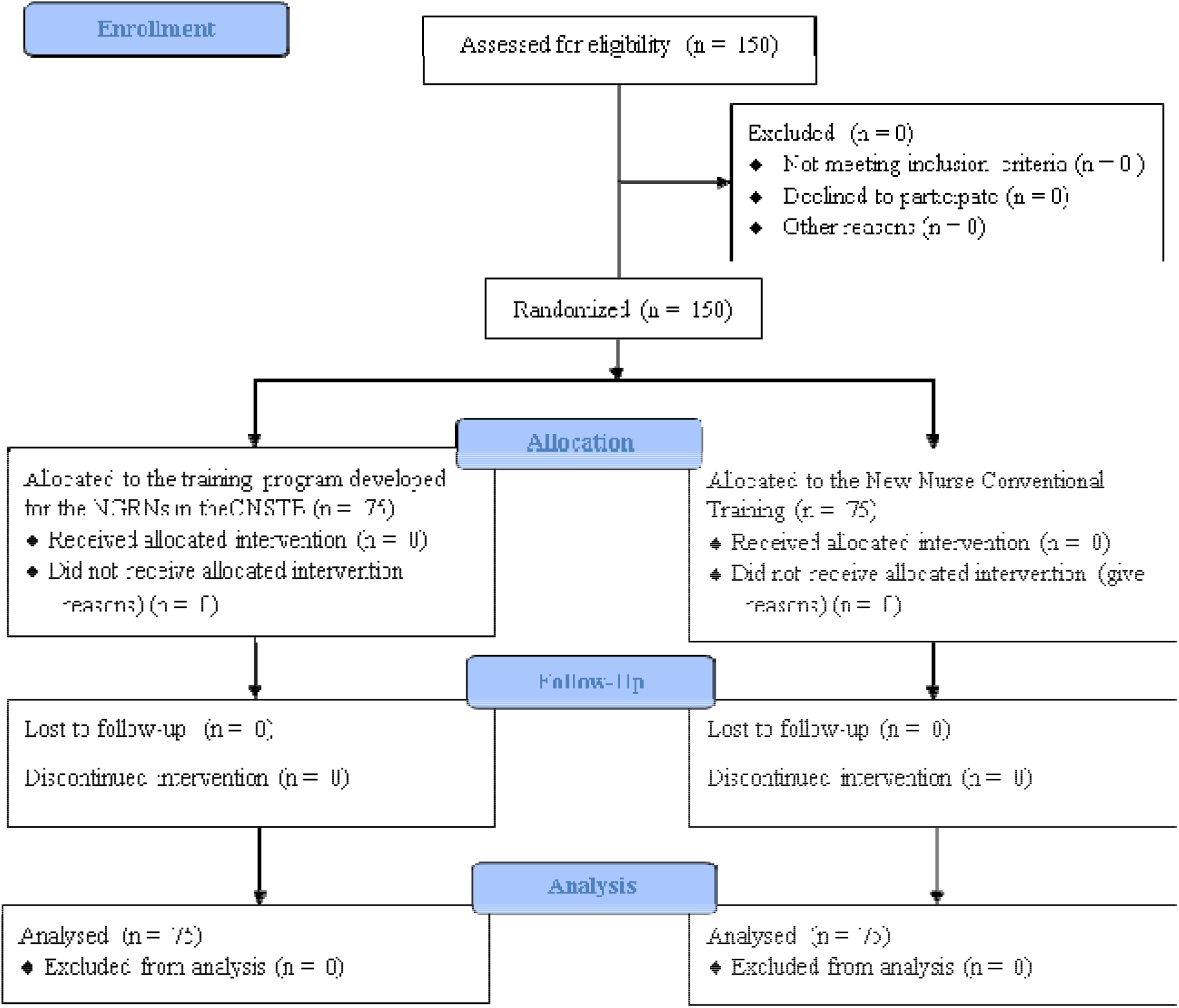
Flow diagram of enrollment, assignment, allocation, follow-up, and analysis of the participants

### Control group

Control group took the New Nurse Conventional Training (NNCT), which included both theoretical knowledge learning and operational skills training. Theoretical knowledge training focused on quality vocation education, interpersonal communication, and occupational safety. At the same time, medical law and ethics training were also included to improve the NGRNs’ legal and ethical awareness. A standardized training manual, training courseware, and a manual of standard nursing operational skills were available for every NGRNs to let them have further self-learning.

### Research group

The training contents of the research group were based on the New Nurse Training Outline (Trial) and Chinese Registered Nurse Core Competency Scale issued by the Chinese State Health and Family Planning Commission [15], and main training courses were designed according to the characteristics of NGRNs (Table 1).

**Table 1.** The training program developed for the NGRNs

| Week | Contents of training program |
| --- | --- |
| 1 | Hospital introduction, organizational structure, rules and regulations, patient safety, communication skills, laws and ethics, infection control |
| 2 | Introduction of nursing work, nursing system & standard, specialized nursing knowledge, work procedure, basic knowledge of scientific research, nursing documents, health education |
| 3 and 4 | Nursing operational skills |

NGRNs, new graduate registered nurses.

In addition to the contents adopted in the control group, the NGRNs in the research group were also required to answer specific clinical questions through clinical practice and literature reading during the operational skills training stage in the third and fourth weeks. The clinical practice was carried out in the CNSTB every workday from 8:00 a.m. to 10:00 a.m. Firstly, the NGRNs observed and exercised the operations that the clinical nursing teachers performed. Secondly, the nurses would try to answer some clinical questions according to their observation and exercise. After the clinical practice, they would spend one hour to have a group discussion and Q&A session about the theoretical and/or clinical issues that they encountered during practices. After 11:00 a.m., the research group and control group would have nursing practices together in the training centre.

### Chinese Registered Nurse Core Competency Scale

At the end of the 4-week pre-job training, the NGRNs’ theoretical knowledge and operational skills were evaluated using a self-assessment questionnaire (each area was scored between 1 and 100), and their core competence levels were assessed using the Chinese Registered Nurse Core Competency Scale.[15] The scale includes 58 items of seven domains, i.e. education and consultation, clinical nursing, leadership, interpersonal relationship, legal and ethical awareness, occupational development, and critical thinking and scientific research. Each item is scored using five-point Likert scale from the lowest competence (0 point) to the highest competence (4 points), with a higher score indicating a higher competence. The internal consistency (Cronbach’s α = 0.89) and criterion validity (r = 0.44) had been examined in previous studies [16].

Except for the self-assessment questionnaire and the scale, we also took notes during the NGRNs’ theoretical knowledge training, wrote reflection diary, and recorded the NGRNs’ scores of the operational skills that were assessed in the CNSTB.

### Statistical analysis

The continuous data were summarized using mean and standard deviation (SD), and categorical data were summarized using percentage. In view of the normal distribution of the data, the differences in the nursing theoretical knowledge and operational skill scores between the groups were tested using the Student’s t-test. A two-sided p-value < 0.05 is considered statistically significant. All the analyses were conducted in the statistical software SPSS version 19.0 (IBM, Armonk, New York).

## Results

The majority (95.3%) of the 150 recruited NGRNs are female (Table 2), and the average age of them is 21.1 ± 1.3 years.

**Table 2.** Characteristics of the NGRNs

|  | Control group | Research group | p-value |
| --- | --- | --- | --- |
| N | 75 | 75 |  |
| Age (mean ± SD, years) | 21.5±1.2 | 21.6±1.4 | 0.639 |
| Sex (%) |  |  |  |
| Male | 4 (5.3) | 3 (4.0) | 0.699 |
| Female | 71 (94.7) | 72 (96.0) |  |
| Training program | New nurses' conventional training (NNCT) | NNCT+ clinical question answering and literature reviewing |  |

SD, standard deviation; NGRNs, new graduate registered nurses.

The average scores of theoretical knowledge and operational skills in the research group were statistically significantly higher than those in control group, which were 88.4 and 94.8, and 81.7 and 90.3 for research group and control group, respectively (Table 3).

**Table 3.** Theoretical knowledge and operational skills scores (mean ± standard deviation)

|  | Theoretical knowledge | Operational skills |
| --- | --- | --- |
| Control group | 81.71 ± 3.16 | 90.34 ± 3.03 |
| Research group | 88.37 ± 4.37*** | 94.75 ± 2.10*** |

\*\*\* p-value < 0.001

Regarding the specific core competencies’ scores, compared to the control group, the research group had statistically significantly higher average scores in education and consultation (2.47 vs. 2.40), clinical nursing (2.87 vs. 2.62), interpersonal relationship (2.56 vs. 2.43), and critical thinking and scientific research (2.78 vs. 2.61) (Table 4). However, there was no statistically significant difference found in leadership ability, legal and ethical awareness, and occupational development (Table 4). The total score of the core competencies of the research group was statistically significantly higher than that of the control group (156.2 vs. 148.8) (Table 4).

**Table 4.** Scores of core competencies

|  | Control group | Research group |
| --- | --- | --- |
| Education and consultation | 2.40±0.21 | 2.47±0.34<sup>*</sup> |
| Clinical nursing | 2.62±0.35 | 2.87±0.23<sup>***</sup> |
| Leadership | 2.46±0.36 | 2.51±0.45 |
| Interpersonal relationship | 2.43±0.39 | 2.56±0.44<sup>**</sup> |
| Legal and ethical awareness | 2.88±0.47 | 2.98±0.52 |
| Occupational development | 2.54±0.69 | 2.65±0.85 |
| Critical thinking and scientific research | 2.61±0.37 | 2.78±0.39<sup>***</sup> |
| Total score | 148.8 ± 26.25 | 156.24 ± 28.32<sup>*</sup> |

\* p-value < 0.05; \*\* p-value < 0.01, \*\*\* p-value < 0.001

## Discussion

### Benefits of CNSTB based training

Usually, hospitals undertake most of the nurse training tasks, while universities offer part of the courses with flexible schedule. The training forms for NGRNs are quite diverse, including setting up learning files, clinical practice, and/or web-based courses, et al. Although the Newly-employed Nurse Training Program (Trial) formulated by the Chinese State Health and Family Planning Commission has paid high attention to the training and role adaptation of the new nurses, the standardized training for the NGRNs in China is still underdeveloped. In order to improve the quality of NGRN training, the pre-job training and clinical practice implemented in the CNSTBs have been widely adopted in the tertiary and teaching hospitals, where NGRNs are trained before they start clinical practices. To our knowledge, our study is the first quasi-randomized controlled study conducted in China that compared the traditional training mode and the innovative training program for NGRNs. The newly established NGRN training program adopted by our research group has shown trilateral win-win results:

1. To nurses, their ability of evidence-based nursing is improved through reviewing a large number of literatures. Their ability of critical thinking is improved through clinical observation and exercises. Through identifying and solving clinical problems, NGRNs learn how to effectively deal with various problems that they might encounter in their future clinical works. Besides, improvement in the core competencies may ensure the NGRNs to adapt to nursing demands as soon as possible, and provide safe and quality care.
2. To hospitals, because the NGRNs who play an important role in clinical services can adapt to their daily job quickly, the innovative CNSTB-based training program may improve the cost-effectiveness of nursing care and result in socioeconomic benefits.
3. To patients, because the NGRNs who take the CNSTB-based training show better interpersonal relationship and can handle their work with critical thinking (Table 4), the patients may receive better and safer nursing services, feel more comfort during the caring, and therefore have higher satisfaction.

### Limitations

Because the NGRNs did not really practice in the clinics, the clinical nursing teachers had no contact with these new nurses. Therefore, the NGRNs had to conducted self-assessment on their theoretical knowledge and operational skills, and were not evaluated by clinical nursing teachers or trainers. There might be bias in the scores. We plan to use longer follow-up after pre-job training in the future study. The NGRNs will be evaluated by the nursing teacher or other more objective methods when they enter clinical practices. The obtained data will be more reliable and objective.

## Conclusion

The innovative CNSTB-based pre-job training program adopted in our study for NGRNs may significantly improve the training effect and is worthy of extensive implementation in the nursing educational settings similar to those in China.

## Abbreviations

CNSTB: the Clinical Nursing Skills Training Base;
DHR: department of human resources;
DN: department of nursing;
NGRN: New graduate registered nurse;
NNCT: new nurse conventional training;
RN: registered nurse;
SD: standard deviation;
TREND: Transparent Reporting of Evaluations with Nonrandomized Designs;
U.K.: the United Kingdom;
U.S.: the United States;

## Practice Points

1. In China, NGRNs face great challenge during the transition from the school to clinical.
2. NGRNs need support and training to speed their transition from new graduates to qualified practitioners, and improve their retention.
3. The innovative training program adopted in the study may significantly improve the NGRNs’ theoretical knowledge, operation skills, and core competencies.

## Declarations

## Ethics approval and consent to participate

The study was approved by the Ethical Committee of the First People’s Hospital of Lianyungang, the First Affiliated Hospital of Kangda College of Nanjing Medical University, China (approval number: KY20171101001). Written informed consent was obtained from all participants.

## Consent for publication

All data were obtained after written informed consent to answer and for data to be analysed and published.

## Competing interests

The authors declare that they have no competing interest.

## Funding

The Authors F.X., H.Z., J.Y., D.L., G.Z., and L.M. were supported by the 13th Five-Year Plan of Education Science of Jiangsu Province, China (20180105-C-b). The funders had no role in study design, data collection and analysis, decision to publish, or preparation of the manuscript.

## Authors’ Contributions

Conceptualization, F.X. and H.Z.; methodology, Y.C.; software; G.Z; validation, Y.W.; formal analysis, G.Z. and Y.X.; investigation, F.X., H.Z., J.Y., and D.L.; funding acquisition, F.X.; resources, F.X. and H.Z.; data curation, F.X. and G.Z.; writing – original draft preparation, F.X. and Y.C.; writing – review and editing, Y.W., H.Z., J.Y., D.L., and L.M.; supervision, H.Z. and Y.C.; projection administration, F.X. and H.Z. All authors have read and approved the manuscript

## Availability of data and materials

The data used to support the findings of this study are available upon request from the corresponding author Hailin Zhang, who may be contacted by email at.

## Acknowledgements

None are to be done.

## Notes

### Competing Interest Statement

The authors have declared no competing interest.

### Author Declarations

The study was approved by the Ethical Committee of the First People's Hospital of Lianyungang (approval number: KY20171101001).

